# Cognitively Healthy Centenarian Brains resist Tau Accumulation and exhibit lower APOE, early proteasome response and maintained β-oxidation

**DOI:** 10.64898/2026.01.08.26343581

**Authors:** Marc Hulsman, Andrea B. Ganz, Meng Zhang, Frank Koopmans, Ka Wan Li, Suzanne S.M. Miedema, Susan K. Rohde, Maruelle C. Luimes, Tjado H.J. Morrema, Annemieke J.M. Rozemuller, Philip Scheltens, Jeroen J.M. Hoozemans, Marcel J.T. Reinders, August B. Smit, Henne Holstege

**Affiliations:** Section Genomics of Neurodegenerative Diseases and Aging, Department of Clinical Genetics, Amsterdam Neuroscience, Amsterdam UMC, Amsterdam, The Netherlands; Department of Molecular and Cellular Neurobiology, Center for Neurogenomics and Cognitive Research, Amsterdam Neuroscience, Vrije Universiteit Amsterdam, Amsterdam, The Netherlands; Alzheimer Center Amsterdam, Department of Neurology, Amsterdam Neuroscience, Amsterdam UMC, Amsterdam, The Netherlands; Department of Pathology, Amsterdam Neuroscience, Amsterdam UMC, Amsterdam, The Netherlands; Delft Bioinformatics Lab, Delft University of Technology, Delft, The Netherlands; VIB-KU Leuven Center for Brain & Disease Research, Leuven, Belgium

**Author notes:** co-first authors (Marc Hulsman, Andrea B. Ganz, Meng Zhang). co-last authors (August B. Smit, Henne Holstege). Corresponding author: Henne Holstege, Amsterdam UMC, The Netherlands.

## Abstract

Brains from cognitively intact centenarians offer a unique window into mechanisms of neuroprotection. We quantified 3,448 proteins by LC–MS/MS in middle temporal gyrus (MTG) from 88 Alzheimer’s disease (AD) patients, 53 controls (50–99 years) and 49 centenarians (100+). After adjustment for cell type composition, Aβ abundance associated with only five proteins, revealing upregulation of HTRA1, LRP1 and NRXN1 as early AD-associated changes. Conversely, tau abundance associated with ∼20% of the quantified proteome, revealing GPRIN1 as the top tau-associated protein. One-third of centenarians had high Aβ abundance, often with Braak neurofibrillary tangle stages IV/V, yet all maintained low local tau abundance. Centenarians showed maintained proteostasis (low ubiquitin peptides, high proteasome components), high PCSK1 (prohormone maturation), high PFKFB2 (glycolysis regulation), mitochondria-coupled lipid β-oxidation and lower APOE levels. Together, these findings suggest a model of neuroprotection in which Aβ facilitates tau seeding, and where proteostatic and metabolic adaptations buffer against additional tau accumulation in centenarians.

## Introduction

Increased life expectancy is accompanied by an increased prevalence of age-related diseases, including Alzheimer’s disease (AD)^1^. The neuropathological hallmarks of AD, extracellular amyloid-β (Aβ) plaques and intracellular neurofibrillary tangles (NFTs), accumulate progressively with age and are present in most individuals aged 90 years and older^2^. Notably, AD-associated pathology is also observed, to variable degrees, in cognitively healthy elderly individuals^3–5^, suggesting a dependency on biological mechanisms that confer resistance (avoiding pathology) or resilience (preserving function despite pathology)^6^.

Cognitively healthy centenarians offer a unique opportunity to uncover endogenous mechanisms that sustain cognitive function despite extreme age. Although around 75% of centenarians develop dementia, about 25% retain cognitive abilities ranging from mild impairment to fully intact performance^7^. In the 100-plus Study, centenarians self-reporting cognitive health at enrollment are longitudinally followed, and one-third consents to brain donation^8^. At donation, participants span a continuum from preserved to declined cognition (**Figure 1B**)^9^. Some centenarians maintained cognitive health despite substantial AD pathology, whereas others showed very limited pathology, suggesting the presence of **resilience** and **resistance** factors, respectively^2,10,11^.

**Figure 1:**
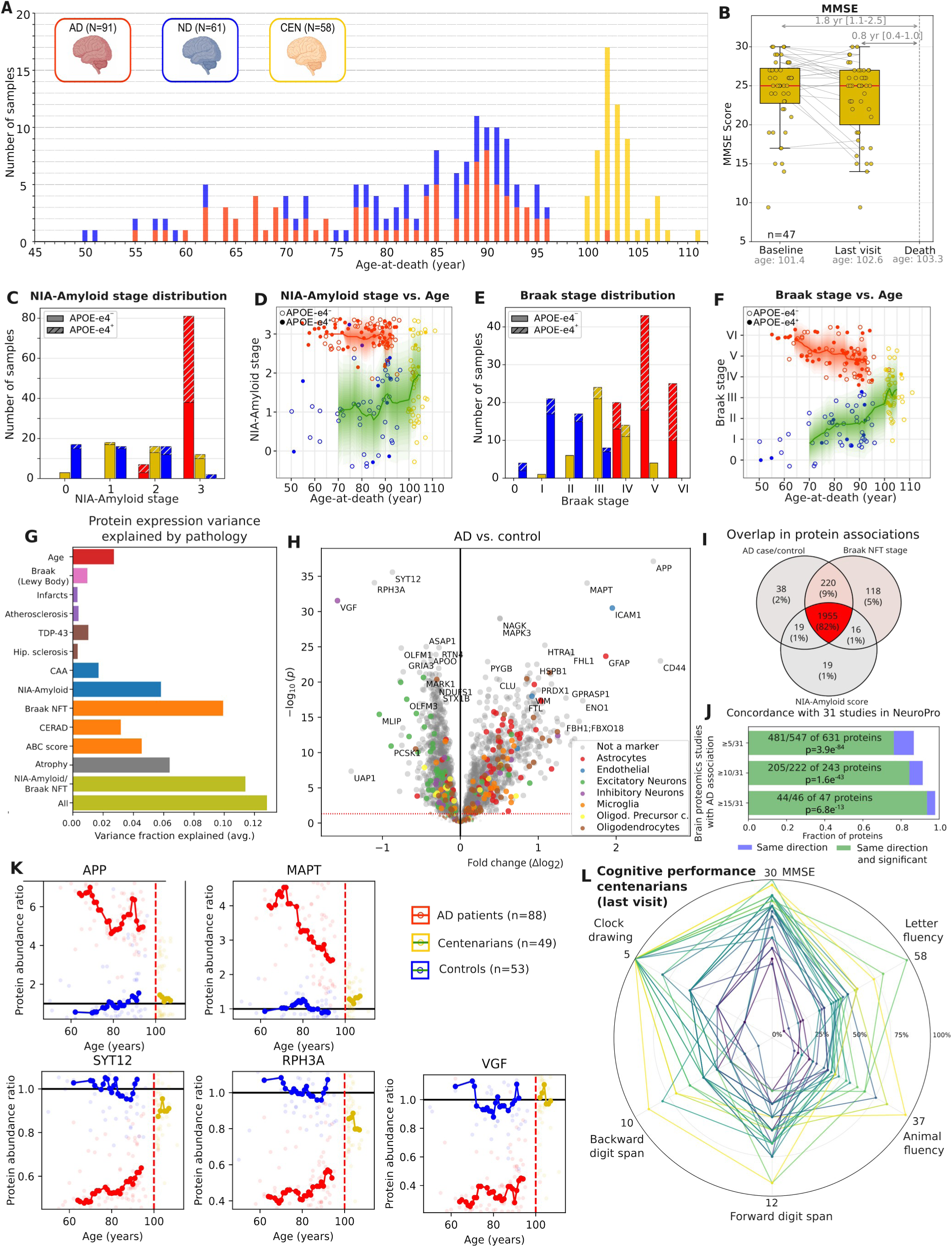
Large-scale proteomic analysis identified AD- and aging-related proteins. **A)** Age distribution of brain-proteomic samples. Red: AD patients (N=88); blue: non-demented controls (N=53); yellow: centenarians (N=49). **B)** The distribution of MMSE scores in centenarians, at baseline and last available visit before death. Trend lines are shown between visits. For n=19 centenarians, baseline = last available visit (no trend line shown). Median time (IQR) to death is shown across the range lines. **C-F)** The number of samples by NIA-Amyloid score (**C,D**) and Braak stage (**E,F**), and the distribution across age, stratified by sample group in samples remaining after QC. Filled circles: carriers of at least one APOE-ε4 allele. **G)** Variance in protein abundance explained by different pathologies and age, with pathologies grouped and colored based on mechanistic similarities. **H)** Changes in protein expression in middle temporal gyrus (MTG) in brains from AD patients and nondemented controls, corrected for age, sex and post-mortem delay; colors: cell type-specific proteins (see methods). **I)** Overlap between proteins that significantly associated with (1) being an AD case relative to controls, (2) increasing Braak NFT stage and (3) increasing NIA-Amyloid score (FDR < 0.05). **J)** Concordance of AD case/control associations and their directions (i.e. increased or decreased with AD) with the significant proteins in at least 5, 10 or 15 of the 31 studies available in the NeuroPro database^19^. Counts of proteins that are significant and have the same direction, and proteins with only the same direction compared to the total number of overlapping proteins. The p-value indicates the significance of the overlap in direction, evaluated using a binomial test. **K)** Age trend of the 5 proteins with the strongest association with AD case/control status. Trend lines are based on a moving average (see methods). Color: type of pathology (e.g. vascular, late, amyloid, tangles). **L)** Cognitive performances of centenarians, last available before death, scaled by maximum (attainable) performance. Each line represents single individual. Lines colored according to average scaled performance. The neuropathological features of this cohort are further described in the Supplementary Data, and available in **Table S1**.

Disentangling the molecular basis of these mechanisms is challenging, as disease- and age-related alterations often overlap^12^. To distinguish these effects, we used LC–MS/MS proteomics to analyze the soluble fraction of brain tissue from AD patients and cognitively healthy individuals spanning an age range covering 50–99 years, and we extended this continuum with centenarians from the 100-plus Study (**Figure 1A**). We focused on the middle temporal gyrus (MTG), a brain region in which Aβ and tau pathologies intersect: amyloid plaques reach the MTG during Thal Phase 2 (preclinical/MCI)^13^, whereas tau tangles affect this region by Braak Stage IV (early AD)^14–16^.

We hypothesized that cognitively healthy centenarians display distinct proteomic signatures compared to AD patients and younger controls, reflecting biological processes that confer long-term resistance or resilience to neurodegeneration.

## Results

### Cohort characteristics and effects of neuropathology

We used LC–MS/MS to quantify proteins in the soluble fraction of laser-captured MTG grey matter from 210 brains. After quality control, 3,448 proteins were quantified in 190 samples (age range 50-111, 74% female): 88 AD patients, 53 non-demented controls (all pathology-confirmed), and 49 centenarians who had self-reported cognitive health at study inclusion, which was confirmed by a proxy (**Figure 1A**). Objective cognitive testing at baseline was available for 47 centenarians, showing that 17% (8/47) had a Mini-Mental State Examination (MMSE) ≤ 20, while 49% (23/47) scored ≥ 26. Brain donors were followed twice yearly until death (median 1.8 years follow-up; IQR 1.0–2.5 years). At the last neuropsychological assessment (median 0.8 years before death, IQR 0.4–1.0 years), 25% scored ≤ 20 and 38% ≥ 26 on the MMSE (**Figure 1B**), indicating that the centenarian cohort captures a broad continuum of cognitive health (**Figure 1L**), enabling analysis of molecular correlates of preserved versus declining cognition.

Neuropathological assessment confirmed that NIA-Amyloid scores^17^ and Braak-NFT stages^18^ were highest in AD patients and increased with age in controls, while centenarians spanned the full range observed in both groups (**Figures 1C–F, S3**). Cerebral amyloid angiopathy, atherosclerosis, hippocampal sclerosis, and infarcts increased with age in both AD and control groups, whereas scores for neuritic plaques, cortical atrophy, and amygdala TDP-43 pathology were elevated specifically in AD patients (**Figure S3**). Lewy body pathology was rare and observed only in some of the oldest individuals. Age, Braak-NFT stage, and NIA-Amyloid score explained the largest variance in protein abundances in this cohort (**Figures 1G, S5**). Epidemiological and neuropathological characteristics are summarized in **Table S1**.

### High concordance of AD-associated proteomic changes across phenotypes and studies

After adjusting for age, sex, and post-mortem delay (PMD), the abundance of 2,232 of 3,448 quantified proteins differed significantly between AD and control brains (FDR < 0.05). The largest increases were observed for microtubule-associated protein tau (MAPT) and amyloid precursor protein (APP) (**Figure 1H, 1K**). Gene Set Enrichment Analysis showed broad downregulation of mitochondrial and synaptic proteins, and upregulation of proteins involved in innate immunity and proteasome activity, with the top enrichment cluster containing the KEGG Alzheimer’s disease pathway (**Table S13**).

Protein abundance changes linked to NIA-Amyloid scores or Braak-NFT stages overlapped by ≥ 96% with those associated with AD diagnosis (**Figure 1I, Table S3**). These protein associations were highly concordant with findings from large independent proteomic resources (the NeuroPro meta-database^19^ and Johnson et al.^12^, **Figure 1J, Sup. Results**) despite methodological and brain-region differences, confirming that the MTG proteome robustly recapitulates canonical AD-associated signatures.

### Centenarians are resistant to Aβ-linked tau accumulation

The abundance of peptides representing the Aβ sequence within APP and the microtubule-binding region (MTBR) region of tau correlated strongly with quantitative IHC for Aβ_40_, Aβ_42_, and pretangles (antibodies G2-10, G2-11 and AT8) and with NIA-amyloid and Braak-NFT neuropathological stages (**Figures 2A–C, S6**). For most proteins, variation in abundance across was better explained by these peptide-derived measures than the neuropathology staging (**Sup. Results**). Therefore, we used the abundance of APP-Aβ and MTBR-tau peptide abundances as measured by LC-MS/MS, hereafter referred to as *Aβ* and *tau,* as primary variables in subsequent analyses.

**Figure 2:**
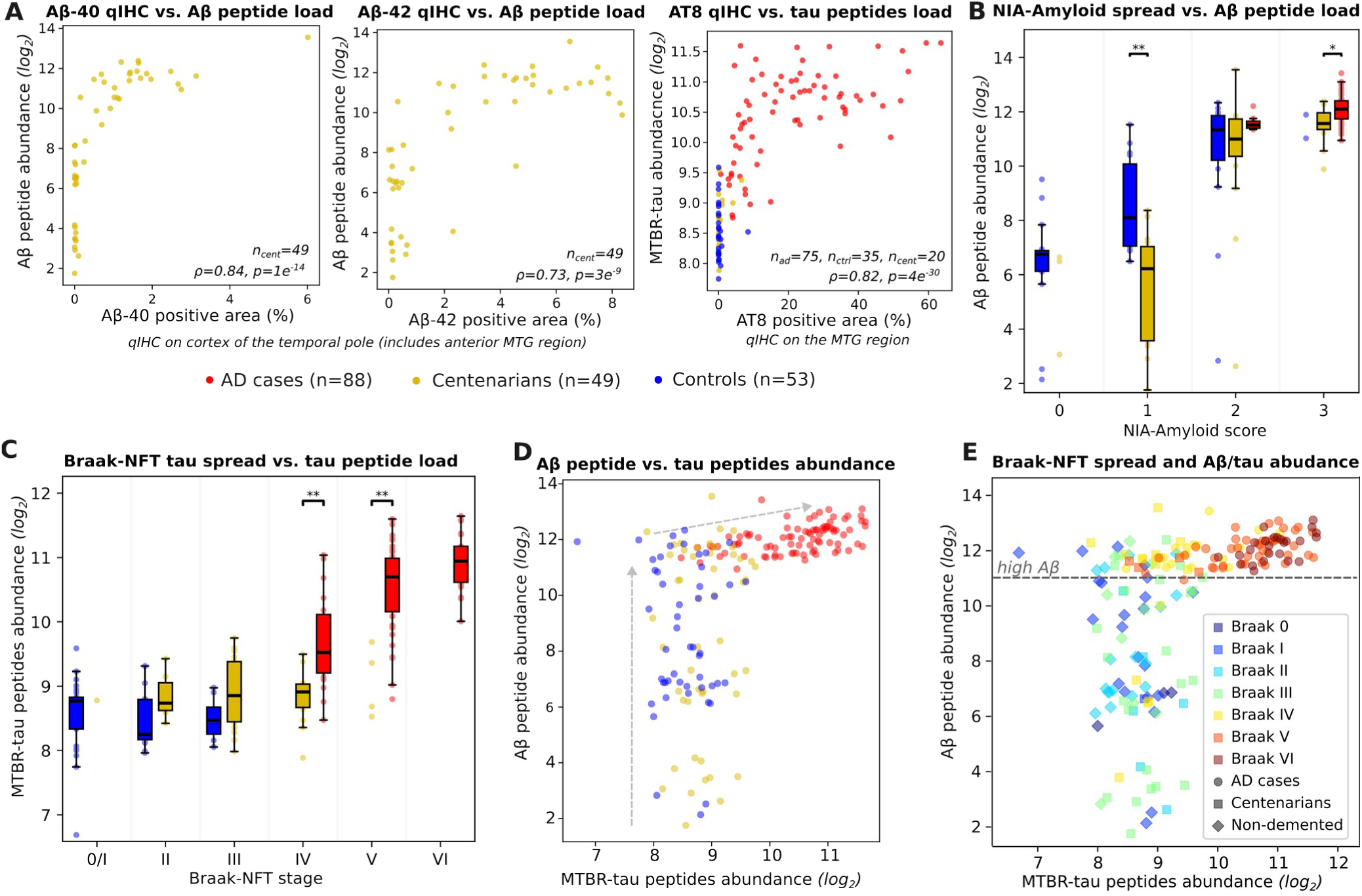
**A)** Comparison in centenarians between Aβ-peptide proteomic measurements (covering amino acids 17-28 of Aβ) and qIHC measurements of Aβ_40_ and Aβ_42_ (antibodies G2-10 and G2-11), on the cortex of the temporal pole (Brodman area 38, adjacent to the MTG). Comparison in AD cases and controls of the average abundance of MAPT peptides covering the microtubule-binding region (MTBR) versus qIHC measurements of AT-8 on the MTG region. AT-8 targets n-terminal MAPT phosphorylation sites known to associate with tau aggregation. qIHC measurements represent the fraction of the positive area (see methods). **B**) Abundance of the the Aβ-peptide, stratified by NIA-Amyloid score and sample group. **C)** Boxplot of peptides covering the MAPT C-terminal MicroTubule Binding Region (MTBR), stratified by Braak stage and sample group. Only 4 controls had Braak 0, and did not significantly differ from controls with Braak I (t-test 0.43). Box plots were omitted for <5 data points. **D)** Aβ and MTBR-tau peptide abundances in the MTG region by sample group. **E)** Abundances of Aβ and MTBR-tau peptides, colored by Braak stage. Horizontal line reflects the threshold for high APP-Aβ used to evaluate tau resistance (Aβ log_2_ abundance=11.0), and is based on the lower limit of Aβ observed in AD cases. Significance: Mann-Whitney U, * (FDR < 0.05), ** (FDR < 0.01), *** (FDR < 0.001).

All AD patients accumulated *Aβ* to the highest levels in the MTG and uniquely showed increased *tau* abundance (**Figure 2D**), in concordance with the amyloid cascade hypothesis stating that amyloid pathology precedes and accelerates tau aggregation^20–23^. Though a third of the centenarians (18/49) reached comparable Aβ levels to AD patients, they maintained control-level tau abundance.

Intriguingly, 13/18 centenarians with AD-like levels of Aβ showed substantial tau *spreading* (Braak IV –V) without increased MTG tau *abundance* (**Table S22**). Across AD patients and controls, Aβ strongly correlated with Braak-NFT stages (r=0.65, p=4.8e^-18^) and with MTG tau abundance (r=0.73, p=1.8e^-24^), and Braak-NFT stages strongly correlated with tau abundance (r=0.82, p=1.8e^-34^). In centenarians, Aβ also correlated with Braak-NFT stages (r=0.52, p=1.0e^-4^) but Aβ *did not* correlate with (MTG) tau abundance (r=0.11, p=0.47), and Braak-NFT stages also did not correlate with (MTG) tau abundance(r_cent_=0.06, p_cent_=0.67). Tau abundances were particularly lower than expected for individuals with Braak stage IV or V relative to AD cases, consistent with these being the stage at which tau reaches the MTG (**Figure 2C, Sup. Results**). This suggests that while Aβ was still linked to the *spreading* of tau pathology in these centenarians, this process did not yet escalate into progressive increase of tau abundance in the MTG.

### Tau, but not Aβ, associates with a large shift in cell composition that is attenuated in centenarians

To dissect independent effects of Aβ and tau abundance on global protein abundance, we fitted a joint model (JM) including both Aβ and tau as predictors while adjusting for covariates sex, age, PMD, atherosclerosis, TDP-43, hippocampal sclerosis and infarcts. Only five proteins, HTRA1, NRXN1, LRP1, ICAM1, and CD44 were significantly associated with Aβ abundance. In contrast, 1,682 proteins were associated with increased tau abundance (49% of the quantified proteome). Applying a JM on neuropathological staging yielded similar results, as only LRP1 correlated with NIA–Amyloid scores whereas 1,310 proteins correlated with Braak–NFT stage (**Figure 3A, Tables S5-S6**).

**Figure 3:**
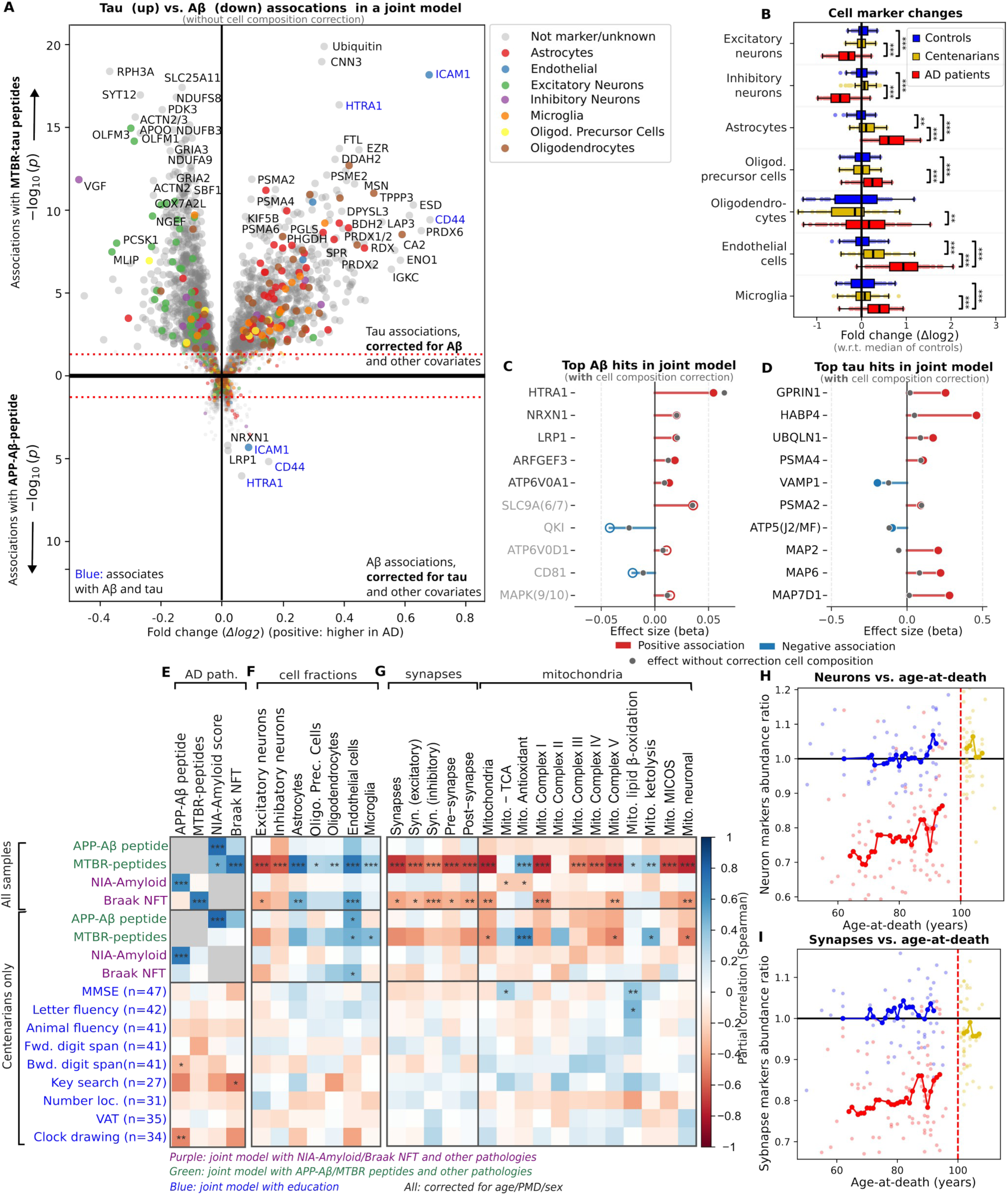
**A)** Protein-pathology relationships were assessed using a joint statistical model (JM) incorporating Aβ and tau levels, while adjusting for sex, postmortem delay (PMD), age, atherosclerosis, TDP-43 (amygdala), hippocampal sclerosis, and infarcts. The upper panel depicts associations with tau abundance, while the lower panel illustrates protein associations with Aβ abundance. The JM accounts for interdependencies between pathologies; for example, associations with Aβ were adjusted for tau levels and other covariates, and vice versa. Proteins that significantly associated with both Aβ and tau are highlighted in blue. **B)** Shared abundance changes of cell-type marker proteins as measured by the first PCA component. Mann-Whitney U test, * (FDR < 0.05), ** (FDR < 0.01), *** (FDR < 0.001). **C,D)** Cell composition correction affects top associatons. Top 10 proteins associated with Aβ and tau after correction for cell composition, age and other pathologies. Non-significant hits (FDR > 0.05) are indicated using a light-gray label. **E-G)** Partial correlations (indicated by color) and regression associations (FDR: * < 0.1, ** < 0.05, *** < 0.01) of AD pathology and cognition measures, with AD pathology measures and measures of relative cell, synapse and mitochondria abundance. Corrected for other pathologies (in case of AD pathology) and education (in case of cognitive measures), as well as age, PMD and sex. Separately for all samples and for centenarians only. **H,I)** Neuron and synapse marker profiles vs. age. Colors indicate AD, centenarian and control cases, according to legend in panel B. Trend lines based on moving average (methods). Ratio’s w.r.t. to controls age 60-70.

We next evaluated cell-type, synaptic and mitochondrial changes using curated marker sets comprising 301 cell-type markers, 44 synaptic markers, and 105 mitochondrial markers (**Methods; Tables S17–S19**). After correction for tau and covariates, Aβ abundance showed only trends toward lower mitochondrial, inhibitory-neuronal, and pre-synaptic markers (**Figures 3F,G**). Although non-significant, these trends align with previous reports^24–28^. In contrast, after correction for Aβ and covariates higher tau abundance was strongly associated with decreased neuron-specific, synaptic, and mitochondrial markers, and increased astrocytic and endothelial markers (**Figure 3B, F**) reflecting glial responses to pathology^29^.

Consistent with their limited tau abundance (**Figure 2D**), centenarians did not show differences in neuronal marker levels relative to controls (p=0.49; **Figure 3H**). Nevertheless, astrocytic markers were elevated (**Figure 3B**) and tau abundance modestly correlated with endothelial and microglial markers (**Figure 3F**), suggesting early glial responses to pathology. Correspondingly, centenarians showed a limited decline in synaptic proteins by 4% (t-test p=0.03, **Figure 3I**), compared to a 18% average reduction in AD patients, consistent with earlier reports^30^, and a more substantial 6% (t-test p=0.003) decline in mitochondrial markers, compared to a 23% reduction in AD patients.

### Aβ abundance associates with executive performance in centenarians

In line with previous findings in 95 centenarians^11^, Aβ abundance was associated with poorer performance on the Clock Drawing Test (CDT, n=34) and Digit Span Backwards (DSB, n=41), whereas the association with the Key Search Test (KST, n=27) did not reach significance (**Figure 3E,4F**). Tau spreading, measured by Braak-NFT stage, showed only trend level association with cognitive tests performance and tau abundance did not associate with test scores, likely reflecting that MTG tau abundances in centenarians largely remained at control levels (**Figure 2D**, **Table S21**). In line with this, we also did not observe any associations with changes in cellular fractions **(Figure 3F)**. Together, our results support a model in which Aβ toxicity sensitizes the brain for tau-spreading and initial cognitive decline, while the centenarians in this study resisted the extensive tau-related cognitive decline that defines end-stage AD.

**Figure 4:**
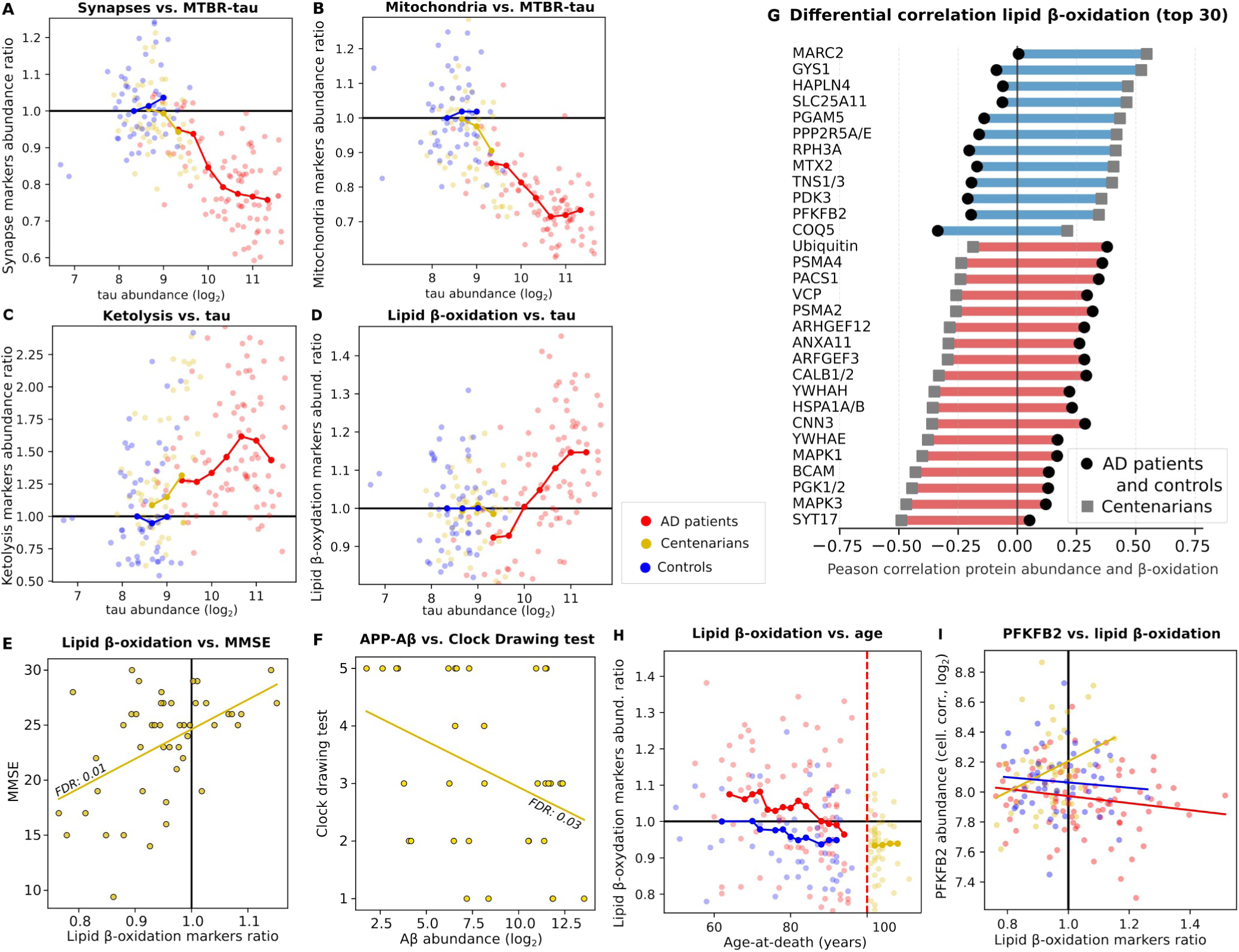
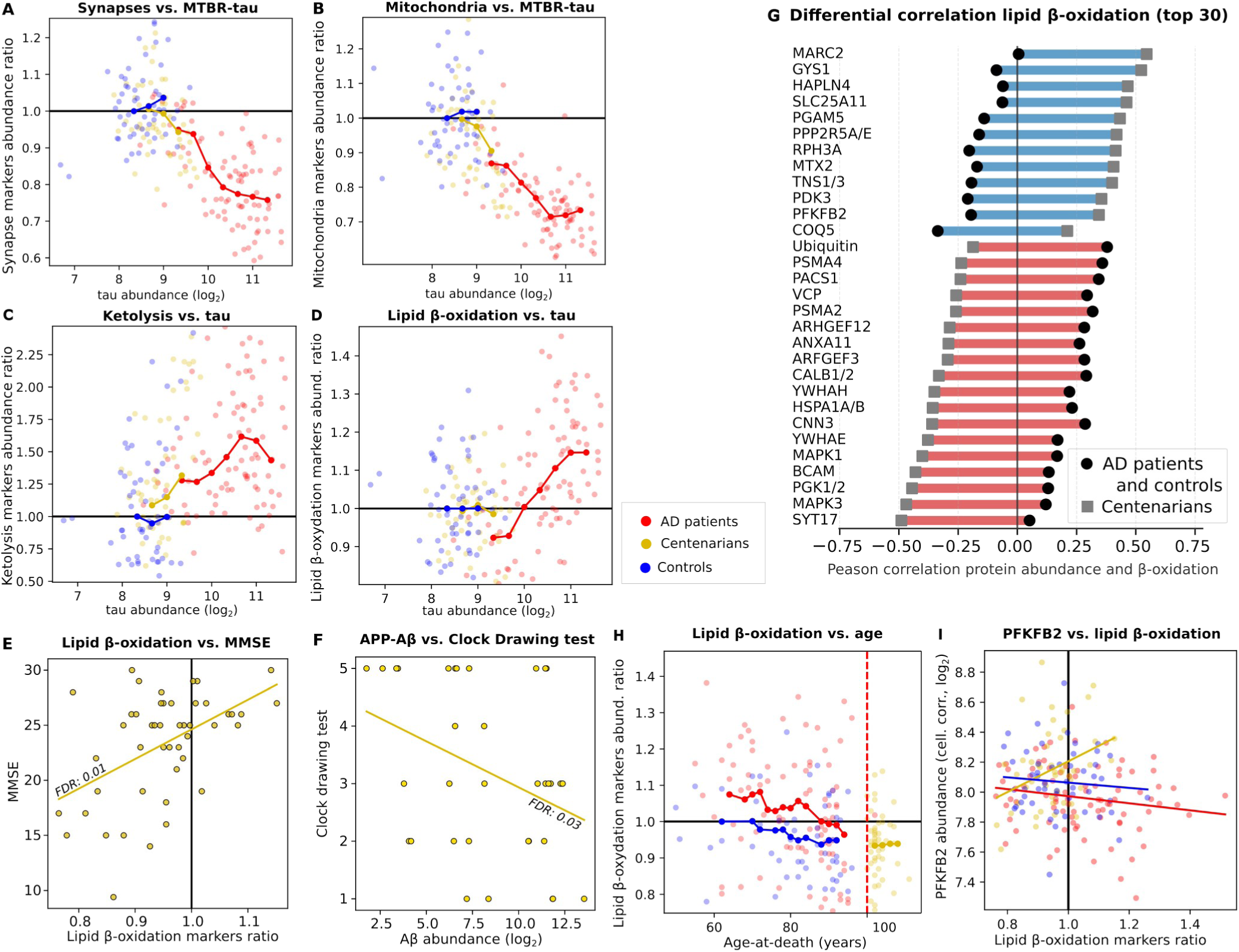
**A-D)** Synapse and mitochondria, ketolysis and lipid β-oxidation marker profiles vs. tau peptides abundance. Trend lines based on moving average (methods). Ratio’s w.r.t. to controls age 60-70. **E,F)** Performance on the Mini Mental State Examination (MMSE) and Clock Drawing Test vs. respectively lipid β-oxidation and Aβ abundance. Trend line based on linear regression. **G)** Top 30 proteins, after exclusion of mitochondrial markers (n=22), with the strongest differences in correlations with β-oxidation between centenarians and AD patients/controls (blue: centenarians have a higher correlation; red: AD and control samples have a higher correlation). **H)** Lipid β-oxidation vs. age-at-death. **I)** PFKFB2 abundance (after cell composition correction) vs. β-oxidation. Trends indicated with linear regression lines.

### Proteins associated with Aβ after adjusting for cell type composition are part of the plaque proteome

After additional adjustment for cell-type composition in the joint model (JM-C model, **Methods**), HTRA1, NRXN1, and LRP1 remained associated with Aβ, while ARFGEF3 and ATP6V0A1 newly emerged as Aβ-associated proteins (**Figure 3C; Table S5**). Notably, the first four proteins were among 48 previously identified plaque proteome proteins^31^. Conversely, the associations between increased Aβ and ICAM1 and CD44 were lost following cell-type adjustment, indicating that their associations were at least partially driven by early amyloidosis-related increases of endothelial cells and reactive astrocytes^32–34^. Functional enrichment revealed that increased Aβ associated with a decrease in G-protein components, PKC targets, and cytoskeletal/mechanosensing pathway components (**Table S14**).

### Cellular composition correction uncovers proteins associated with tau abundance hidden by neuronal loss

Adjusting for changes in cell type composition retained 494 of the tau-associated proteins, and revealed 174 additional proteins, reflecting ∼20% of the considered proteome (**Figure 3D, Table S6**). Normally masked by neuronal cell loss, neuronal upregulation of GPRIN1 emerged as top tau-associated event. GPRIN1 is a signaling adaptor protein that promotes neurite outgrowth by linking G-protein signaling to actin cytoskeleton remodeling, including the recruitment of the α7 nicotinic acetylcholine receptor (α7-nAChR) to growth cone membranes during brain development^35^. If GPRIN1 similarly regulates α7-nAChR membrane expression in the Alzheimer’s disease brain, this would be noteworthy, as α7-nAChR has a high affinity for Aβ. Through receptor-mediated endocytosis, subsequent activation of GSK-3β, and ultimately tau phosphorylation, α7-nAChR may contribute to Aβ-driven neurotoxicity^36–39^. Other unmasked top-upregulated proteins included microtubule-associated proteins MAP2, MAP6, MAP7D1 and stress-granule associated protein HABP4. Functional enrichment of tau-associated proteins revealed 31 clusters: 29 downregulated clusters represented predominantly mitochondrial proteins, 2 upregulated clusters representing microtubule binding and the proteasome (**Table S15**). Notably, upregulation of HTRA1 was the top amyloid-associated and 4^th^ strongest tau-associated change, and its abundance correlated most strongly with the abundance of MAPT, APOE, APP and ARFGEF3, reflecting the central role of HTRA1 in AD disease processes^40,41^ (**Table S25**).

### Proteins distinguishing tau spreading, accumulation, and resistance

To distinguish proteins associated with tau spreading from those reflecting tau abundance, we expanded the JM-C model to include NIA-Amyloid and Braak staging scores alongside Aβ and tau peptide measures. After controlling for tau abundance, cell composition, and other pathologies, we identified three proteins specifically associated with Braak stage (tau spread): SYT12, MAPK3, and SLC2A13 (**Table S8**). Notably, SYT12 (negatively associated) has been linked to cognitive performance^42^, while MAPK3 and SLC2A13 (both positively associated) are respectively linked to early tau deposition^43^ and γ-secretase-mediated Aβ production^44^. Conversely, 495 proteins were associated with tau abundance after correction for tau spread (Braak stage). The top ten included microtubule-associated proteins (MAP7D1, MAP2, MAP6), proteasome-related proteins (UBQLN1, PSMA4), synaptic proteins (GPRIN1, SHANK2), and proteins involved in RNA regulation (HABP4, PABPC1) and NMT1. Furthermore, by comparing tau-resistant centenarians and controls (high Aβ/low tau; **Figure 2E**) with AD cases (high Aβ/high tau), revealed three proteins associated with resistance to tau accumulation despite high Aβ: RBMX/RMBXL1 (RNA-binding motif protein X-linked), PPP2R2A, and EGFR (**Table S7).** For a detailed discussion of these proteins, see Supplemental Results.

### Mitochondrial decline accompanied by compensatory lipid metabolism

Tau abundance tracked negatively not only with synaptic markers (r=-0.71; **Figures 3G, 4A**) but also with mitochondrial markers (r=-0.77; **Figures 3G, 4B**). Notably, the decline in mitochondria markers was selective: inner membrane respiratory oxidative phosphorylation machinery (OXPHOS complexes I, III–V) and components of the mitochondrial contact site and cristae organizing system (MICOS) decreased, in line with literature^45^, and consistent with cristae disruption^46,47^. In contrast matrix-localized pathways, including antioxidant defense, the tricarboxylic-acid cycle, complex II (the matrix-facing, inner-membrane enzyme that links the TCA to the ETC), ketolysis, and lipid β-oxidation) were preserved or increased (**Figure 4C,D; Table S21**). These findings suggest that as respiratory capacity at the inner mitochondrial membrane declines, matrix enzymes partially compensate by sustaining acetyl-CoA production and redox buffering through enhanced ketolysis, while astrocytes support energy metabolism with increased ketone-body provision and mitochondrial β-oxidation. Consistent with this astrocytic response, β-oxidation (CPT1A, CPT2, ACADVL, HADHA, HADHB, ACAA2, ACSL1) and ketolysis abundances correlated with astrocyte markers (r=0.60 and 0.56, respectively). However, β-oxidation only weakly correlated with ketolysis (r=0.18): with decreasing mitochondrial-membrane protein levels, ketolysis increased more strongly than β-oxidation (r=-0.62 and -0.34, respectively; **Table S28**)^48^. Conversely, β-oxidation showed a stronger inverse correlation with neuronal markers than ketolysis (r_β-oxid_=-0.55; r_keto_=-0.19), consistent with β-oxidation being relatively enriched in astrocytes, whereas ketolysis is primarily neuronal. A similar pattern, particularly pronounced by elevated antioxidant defenses, was observed in centenarians, indicating that this compensatory metabolic rerouting is already engaged even at low tau levels.

### β-oxidation marks gliosis in AD but preserved metabolism and cognition in centenarians

Intriguingly, centenarians with higher aggregate levels of lipid β-oxidation proteins also showed better cognitive performance (MMSE: r=0.39, FDR=0.01, n=47; Letter Fluency Test (LFT): r=0.45, FDR=0.05, n=42; **Fig. 4E, Table S21**). Individual β-oxidation proteins showed positive but weaker correlations, the aggregate signal was stronger, consistent with a pathway-level effect (**Table S24**). In contrast, ketolysis did not relate to MMSE (r=0.20, FDR=0.68). These observations argue against β-oxidation representing only reactive gliosis or mitochondrial injury and instead support its role as a flexible metabolic adaptation that support neuronal survival and redox homeostasis under healthy and early stress conditions. Indeed, in centenarians, β-oxidation showed a markedly weaker association with astrocyte markers than in AD (r_cent_=0.29 vs r_AD_=0.81) and a positive, rather than negative, correlation with mitochondrial proteins (r_cent_=0.29; r_AD_=-0.42; difference p=1.1e^−5^), consistent with preserved neuronal/mitochondrial metabolism. By contrast, ketolysis remained negatively associated with mitochondrial markers in both groups (r_AD_=-0.50; r_cent_=-0.53).

As many as 442 proteins correlated differentially with β-oxidation between centenarians and AD cases/controls (**Table S27**), reflecting fundamentally distinct regulatory states of energy metabolism. After cell-composition correction, the abundance of 90 proteins remained differentially correlated at an explorative FDR<0.1 (**Table S27**), including 22 mitochondrial OXPHOS markers, consistent with β-oxidation being coupled to respiration in centenarians but decoupled or anti-coupled in AD. Additionally, in centenarians, higher β-oxidation coincided with higher PDK3 abundance (a kinase that inhibits the flux of glycolytic-derived pyruvate into the TCA cycle), reduced levels of glycolytic enzymes (PGK1/2), and increased glycogen synthase 1 (GYS1). This pattern suggests a coordinated rerouting of glucose away from oxidative metabolism towards, glycogen storage or lactate shuttling to support neuron–astrocyte metabolic exchange. In AD, however, β-oxidation correlated with lower PDK3 and higher PGK1/2, indicating a scenario in which activation of β-oxidation and glycolysis, are simultaneously activated, a configuration consistent with metabolic stress rather than controlled adaptation.

These associations further indicate that centenarians preferentially rely on fatty-acid oxidation rather than glycolysis, a configuration that may help mitigate lipotoxic stress. Consistent with this, β-oxidation showed opposing correlations with proteostasis pathways (HSP70 family, ubiquitin–proteasome system, VCP) and tau-regulatory proteins (14-3-3 family YWHAE/YWHAH; MAPK1–3; PP2A regulators PPP2R5A/E; Fig. 4g). Whereas the associations in AD likely reflect activation of stress-response mechanisms, the inverse correlations in centenarians suggest that preserved β-oxidation either reflects or contributes to the avoidance of tau-related stress in this group.

Together, these patterns suggest a state-dependent behavior of β-oxidation: in centenarians, higher β-oxidation aligns with preserved, respiration-coupled metabolism, whereas in established AD it appears as a compensatory, reactive program that emerges alongside mitochondrial injury. This interpretation is supported by the biphasic relationship with tau in **Figure 4D**: centenarians cluster in the low-tau, pre-inflection regime (r=−0.12), while AD brains occupy the post-inflection regime, where β-oxidation increases with tau (r=0.55), marking a shift from physiological support to stress-responsive remodeling.

Notably, the aggregate marker for β-oxidation abundance declined with age-at-death (**Figure 4H**), but it did not achieve significance (p=0.13) in the JM-C model, suggesting that age-related changes were mainly driven by cell composition changes. Beyond β-oxidation, exploration of the 10 top MMSE-associated proteins, though non-significant, highlighted several additional lipid metabolism components (**Table S24**): ACSS1 and MGLL (acetate and triglyceride catabolism) increased with MMSE, whereas ACSL3/4 (directing fatty acids toward anabolic fates) decreased. Their weak correlation with β-oxidation (r=0.31, 0.23 and -0.10 respectively) suggests complementary, partly independent mechanisms that converge on efficient acetyl-CoA generation in a glycolysis-constrained environment.

### Proteins associated with AD pathology progression

Individuals who delay accumulation of pathology to late ages may possess protective mechanisms or lack disease-promoting factors. We therefore hypothesized that proteins associated with both age-at-death and AD pathology may capture biological processes that influence the rate of pathology accumulation. Across the cohort, the joint model (JM-C) identified 946 proteins associated with Aβ or tau, independent of cell-type composition and other pathologies (FDR < 0.1; **Tables S5, S6**). Of these, 46 also varied with age-at-death after correcting for pathology (FDR<0.05; **Table S9**). Twenty-seven proteins showed one of three patterns consistent with involvement in protective or harmful processes: protective, up-with-age-at-death/down-with-pathology (n=3), compensatory, up-with-age-at-death/up-with-pathology (n=18), or harmful, down-with-age-at-death/up-with-pathology (n=5); the remaining proteins followed an age-related decline pattern (down-with-age-at-death/down-with-pathology), characteristic of neurodegeneration (**Fig. 5A; Table S23**). Here, we highlight the six signals for which the abundance in centenarians lies outside the range of both controls and AD cases: APOE, ubiquitin-encoding proteins, PFPKFB2, LRPAP1, SHANK2 and PCSK1. Other proteins are discussed in the Supplementary Results.

**Figure 5:**
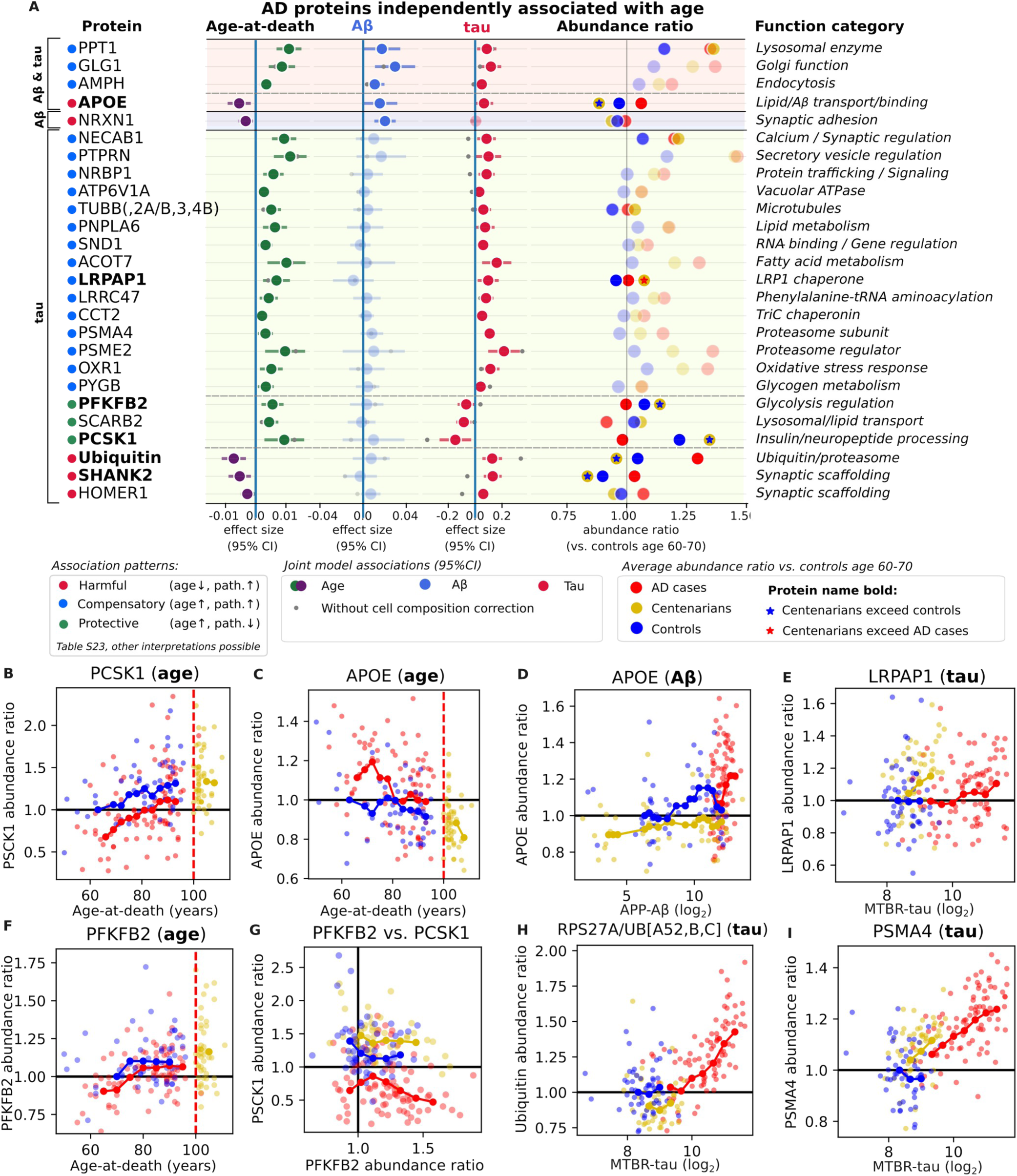
**A)** Proteins of which abundances associated with age-at-death (FDR < 0.05) and independently with Aβ and/or tau abundance (FDR < 0.1) after adjusting for atherosclerosis, TDP-43 (amygdala), hippocampal sclerosis, infarcts, cell-type composition, sex and PMD. Effect sizes without correction for cell composition direction are highlighted with a gray dot. Proteins have been grouped based on the presence of an association with Aβ and/or tau (p < 0.05), and subgroups are delineated based on the directions observed with age and pathology. Within subgroups, proteins are ordered on the strength of the age association. Lack of association with pathology (p > 0.05) is shown with increased transparency of the plot markers. Average abundance ratios relative to ND controls of age 70 (see methods) is shown for the different groups. Proteins for which centenarians had an average abundance between AD patients and controls are shown with increased transparency. Proteins for which centenarians significantly exceeded the AD case and control range in a certain direction are shown in bold and centenarian average abundances are highlighted with a star symbol: blue when exceeding control abundances, red when exceed AD abundances. **B-I)** The abundance of selected proteins with age, Aβ and tau and each other. Protein abundance ratio: all abundances were calculated relative to the abundance in 60- to 70-year-old controls and after correction for cell composition.

#### APOE abundance declines with increasing age-at-death

APOE exhibited a *harmful* pattern, setting the centenarians apart (centenarians < controls < AD cases; p=9.5e⁻^4^). APOE abundance showed modest positive correlations with Aβ and tau pathology (p_aβ_=0.04; p_tau_=0.02; **Figure 5C,D**). However, higher age-at-death was associated with lower APOE levels after correction for pathology and cell composition (FDR=0.04, p=0.001), a finding that remained across multiple sensitivity analyses: when restricting controls and centenarians (p=0.006) and after adjusting for APOE-ε2 or -ε4 genotype (p=0.002). Despite this, ε4 carriers still exhibited higher overall APOE abundance (p=0.002).

APOE’s central role in both amyloid and tau pathology was further highlighted by the top 10 proteins with which its abundance correlated: HTRA1 (r=0.65), CLU (r=0.47), MAPT (r=0.45) and APP (r=0.39) (**Table S25**). Mechanistically, APOE contributes to lipid transport and Aβ clearance but also facilitates aggregation, such that lower levels might limit plaque formation^49–52^. Consistent with its role in lipid transport, its abundance correlated positively with markers of β-oxidation (r=0.42), also in centenarians (r=0.31). These finding support the hypothesis that the capacity to finely balance APOE’s multiple functions, given age and genotype, may contribute to preserved metabolic integrity and maintaining cognitive health.

#### Preserved proteostasis and a distinct ubiquitin–proteasome signature

A second distinguishing feature of centenarian brains was a shift in protein homeostasis components. Peptides tracking ubiquitin, derived from the four ubiquitin-encoding proteins (fusion-proteins RPS27A, UBA52 and poly-ubiquitin-carrying UBB and UBC) reflected a *harmful* pattern, with lower abundance in centenarians than in controls or AD cases (p=3.7e^-3^; **Figure 5H**). In contrast, proteasome subunits PSME2 and PSMA4 increased with both age-at-death and tau pathology (a *compensatory* pattern, **Figure 5I**). AD brains thus showed concurrent elevation of ubiquitin and proteasome proteins, consistent with proteotoxic stress^53^, whereas centenarians displayed the opposite pattern: reduced ubiquitin with higher proteasome abundance, suggesting preserved proteostatic capacity in extreme aging, potentially enabling more efficient substrate turnover before aggregation burdens arise.

#### Metabolic regulation linked to neuronal resilience

Metabolic regulation further distinguished centenarians.

The glycolytic regulator PFKFB2 (**Figure 5F**) showed a protective pattern (centenarians > controls > AD; p=0.036; Figure 5A. Per-cell PFKFB2 abundance associated with maintained OXPHOS (r≥0.36) and higher β-oxidation (r=0.34), alongside lower antioxidant and ketolysis proteins (r=−0.42, −0.33). This constellation reflects efficient respiratory metabolism using both glucose and lipid oxidation rather than reliance on ketones; notably, the PFKFB2–β-oxidation relation reversed in AD (r=−0.18). Mechanistically, PFKFB2 supports metabolic flexibility, by lowering glucose oxidation and increasing fatty-acid oxidation (FAO) when transitioning from fed-to-fasted states; loss of PFKFB2 unleashes glycolysis leading to drops in circulating glucose during fasting^54^.

#### Maintained LRP1 trafficking through LRPAP1

LRPAP1 (centenarians > AD > controls, p = 0.029) displayed a *compensatory* pattern. LRPAP1 is an LDL-receptor chaperonin, which abundance rose with age-at-death and tau, with centenarians showing a more tau-responsive increase compared to AD cases (**Figure 5E**). Although not significant, LRPAP1 showed the second-strongest (positive) association with MMSE (**Table S24**). Intracellular LRPAP1 facilitates LRP1 trafficking to the plasma membrane by preventing premature ligand binding^55^. LRP1, which we identified as one of the five proteins associated with increased Aβ, is an endocytic receptor for APOE, Aβ and tau, and plays an important role in Aβ clearance and has also been linked to tau clearance and spreading^56,57^. LRPAP1 may also be released by activated microglia, where it inhibits extracellular (synaptic) phagocytosis, Aβ uptake and Aβ aggregation^58^. This release might possibly reflect the observed increases with tau.

#### Attenuated synaptic protein response in centenarians

Synaptic remodeling typically accompanies both aging and pathology. Multiple synaptic (UNC13A, PSD2, DLG4/PSD-95) and cytoskeletal proteins (filamins, α-actinins) declined with age and tau (age-related decline pattern, **Table S9, Figure 5A**). However, several synaptic proteins (NRXN1, SHANK2, HOMER1) increased on a per-neuron basis with pathology burden, possibly indicative of a response to Aβ- (NRXN1) or tau- (SHANK2, HOMER1) associated synaptic remodeling. Interestingly, these same proteins showed age-related decreases, particularly in SHANK2 (centenarians < controls < AD cases, p=0.018), suggesting an innate increased activity to downregulate pathology-induced synaptic response in centenarians, reflecting an adaptive strategy that limits synaptic remodeling and prevents progression into maladaptive compensatory states characteristic of AD.

PCSK1, which had a higher abundance in centenarians and showed a *protective* pattern (centenarians > controls > AD cases; p=0.018; **Figure 5B**), and encodes a prohormone convertase essential for the maturation of neuropeptides, e.g. insulin, GLP-1, and neuropeptide Y and others. Per-cell abundance of PCSK1 associated with excitatory and inhibitory neurons marker abundance (r=0.45, 0.33).

### Proteins associated with ageing

Functional enrichment analysis of the age-associated proteome (**Table S16**), obtained using the JM-C model which adjusts for cell-type composition, Aβ and tau abundance, and other covariates, revealed a divergence between upregulated metabolic support systems and downregulated synaptic machinery during aging. Processes significantly upregulated with increasing age were dominated by cellular maintenance and metabolic regulation. The top enriched clusters involved tRNA aminoacylation and aminoacyl-tRNA synthetase activity (Cluster 1; e.g., *NARS, FARSA*), suggesting an increase in translational machinery, alongside ribose phosphate and purine nucleotide biosynthesis (Cluster 9) and glycolytic processes (Cluster 22; e.g., *PFKP, PFKFB2*). Additionally, we observed an upregulation of lysosomal lumen activity (Cluster 12), potentially reflecting increased demands for protein degradation and waste clearance in the aging brain.

Conversely, processes downregulated with age (negative NES) overwhelmingly centered on neuronal communication and synaptic structural plasticity. The most significant declines were observed in glutamate and ion transmembrane transport (Cluster 3; e.g., *SLC1A3, SLC1A2*), suggesting a compensatory effect in reduced excitatory neurotransmitter release (e.g. UNC13A). This was accompanied by a broad loss of enrichment for synaptic plasticity, postsynaptic density organization, and dendritic spine development (Cluster 4; Cluster 6), driven by key postsynaptic scaffolds and receptors such as *SHANK3, DLG4 (PSD-95),* and AMPA-type glutamate *GRIA* subunits. Collectively, these results suggest that aging in the brain, independent of Alzheimer’s pathology, is characterized by a preservation or compensatory ramp-up of basic metabolic functions amidst a progressive decline in (excitatory) synaptic function.

## Discussion

Neuropathological studies indicate that Aβ deposition typically precedes tau spreading beyond the entorhinal cortex, and that tau accumulation is the primary correlate of neurodegeneration and cognitive decline^59–62^. Yet in the oldest-old, the predictive power of Braak staging for cognition diminishes; individuals, even some at Braak IV/V, may retain relatively preserved function^63^. By combining laser-capture proteomics in middle temporal gyrus (MTG) with detailed neuropathological staging and cognition measures, our findings provide a possible basis for this paradox: centenarians may accumulate substantial Aβ and show widespread tau *spread* (as indicated by Braak stage) yet lack the local tau accumulation that is strongly linked to neuronal and synaptic loss.

Using a joint model to disentangle Aβ from tau effects, we showed clear asymmetry. After covariate adjustment, Aβ abundance associated with only five proteins, whereas tau associated with ∼50% of the quantified proteome, reflecting broad loss of neuronal, synaptic, and mitochondrial markers and increases in astrocytic/endothelial signatures. Centenarians diverged from the AD trajectory in two critical aspects. First, one-third reached Aβ levels comparable to AD, but they maintained low local tau abundance in MTG despite frequent Braak IV/V staging. Second, neuronal marker levels were preserved, synaptic protein downregulation was modest (∼4% vs ∼18% in AD), and mitochondrial protein decline was far smaller than in AD, together supporting a decoupling of Aβ from tau *accumulation* and its downstream cellular remodeling in the MTG. These observations align with the amyloid cascade framework^22,23^ but extend it by identifying a distinct centenarian state in which Aβ still relates to *tau spread* yet does not drive the pathological tau escalation in MTG that typifies AD.

Cell-composition-adjusted analyses revealed five proteins associated with Aβ abundance (HTRA1, LRP1, NRXN1, ARFGEF3, ATP6V0A1), of which the first four were identified as part of the plaque proteome^31^. Notably, HTRA1 was also associated with tau-abundance, positioning HTRA1 upregulation as a central event in early AD. HTRA1 has both chaperone and serine protease activities and contributes to extracellular matrix (ECM) remodeling and TGF-β signaling^64^.

After cell-composition-adjustment, tau abundance still associated with abundance-changes in ∼20% of the identified proteome, unmasking tau-linked neuronal factors, notably GPRIN1, a regulator of actin-dependent neurite outgrowth that might couple α7-nAChR:Aβ interactions to kinase activation and tau phosphorylation^35,37–39^.

Extending the model to separate tau *spread* (Braak) from local MTG tau *abundance,* identified spread-specific correlates, notably ERK1/MAPK3 which is activated by Aβ and has been implicated in tau phosphorylation and early tau deposition^65,66^. Conversely, resistance to tau accumulation under high Aβ was characterized by several proteins, most notably downregulation of RBMX/RMBXL1, which is known to inhibit splicing of exon 10 of MAPT, which would shift tau isoforms towards the more aggregation-prone 4R form^67^. RBMX is also detected in the stress granule proteome^68^, a cellular compartment implicated in promoting tau aggregation^69^.

Our observation of low tau accumulation in the centenarians supports a threshold model: whereas Aβ facilitates seeding and propagation, avoiding the transition to self-amplifying locally escalating tau accumulation appears to depend on the integrity of proteostatic and metabolic buffering systems. Several centenarian-specific signatures appear consistent with such buffering.

First, we observe a distinctive ubiquitin-proteasome profile in centenarians: *lower* ubiquitin peptides coupled with *higher* proteasome components (PSME2, PSMA4) suggests efficient pre-aggregate clearance, in contrast with the stress-associated pattern observed in AD (higher ubiquitin alongside elevated proteasome components)^53^.

Second, centenarians predominantly maintained a low-tau state in which higher abundance of β-oxidation-related proteins correlated with better cognitive performance. In these individuals, β-oxidation also remained positively coupled to mitochondrial respiration; by contrast, in AD, β-oxidation was likewise upregulated but tracked with gliosis and inversely with OXPHOS and MICOS components^46,47^ reflecting reactive metabolic stress rather than physiological compensation. This biphasic pattern is consistent with an astrocyte-mediated lipid detoxification and metabolic support program that is initially protective but becomes a stress-driven response as tau pathology advances^48,70,71^. Notably, β-oxidation correlated with several tau-regulatory proteins both in AD and within the centenarians’ low-tau regime, suggesting a direct intersection between lipid metabolic regime and tau regulation.

Maintained flexibility in these metabolic pathways may be a defining feature of centenarian resilience. The importance of this was underscored by the glycolysis regulator PFKFB2, which was elevated in centenarians but lowest in AD, and associated with a metabolic profile characterized by preserved OXPHOS and β-oxidation, together with reduced antioxidant and ketolysis signatures. Against a backdrop of age-related declines in aerobic glycolysis, the centenarian feature of dual fuel capacity may reflect complementary adaptations that maintain energetic flexibility in late life^72,73^.

Finally, APOE increased with Aβ and tau, but decreased with higher age-at-death, with centenarians having the lowest levels. Given APOE’s roles in lipid transport and in both Aβ clearance and aggregation^49,50^, lower levels could reduce plaque seeding, while still supporting efficient lipid handling in a preserved metabolic environment. LRPAP1, an intracellular chaperone that facilitates LRP1 trafficking, was higher in centenarians and increased with age and tau, potentially sustaining LRP1 availability for lipid-cargo processing. In addition, LRPAP1 can also be released extracellularly by activated microglia, where it inhibits phagocytosis, Aβ uptake, and Aβ aggregation^56–58^, suggesting that compartment-specific regulation of LRPAP1 may influence how the ageing brain balances clearance versus containment of pathogenic proteins.

This study has some limitations. First, analyses were confined to a regional sample (MTG), and it remains unclear whether the identified mechanisms generalize to other vulnerable brain regions. We did not specifically search for modified peptides and the depth of analysis may prevent inclusion of data from sparse cell types, (i.e. microglia). Our cell deconvolution is based on cell-type–specific markers derived from transcript expression data, which is known to correlate imperfectly with protein abundance. Our linear modeling does not include potentially non-linear processes. Moreover, we did not explore potential sex-specific effects, which are likely to occur. Furthermore, the centenarians in this study represent a selected subgroup enriched for survival and cognitive resilience, and ascertained specifically to identify protective associations. As a result, the findings may not fully extend to the general centenarian population.

Taken together, our results support a model in which cognitively healthy centenarians exhibit a state of resistance to tau accumulation despite substantial Aβ deposition and tau spread. This resistance appears underpinned by preserved proteostasis and a flexible, mitochondria-coupled lipid metabolic program. These buffering mechanisms may remain functional in exceptionally long-lived individuals and are reflected in several centenarian-specific molecular signatures, including enhanced proteasomal and lysosomal activity, modulation of the APOE/LRP1 axis, and metabolic programs e.g., linked to PFKFB2.

Future work should validate these signatures in more diverse aging cohorts and expand to additional brain regions affected earlier in AD, and incorporate single-cell and epigenomic approaches to resolve regulatory mechanisms across cell types. Ultimately, targeting the metabolic and proteostatic pathways that remain functional in exceptional aging may offer new therapeutic strategies to delay or prevent progression to symptomatic AD.

## Data Availability

All data produced in the present study are available upon reasonable request to the authors

## Acknowledgements

We thank and acknowledge all participating centenarians and their family members and the team who recruited and visited the centenarians and their family members over the years, who collected the neuropsychological data and informed them about the option for brain donation: Chandeny Bennewitz, Debbie Horsten, Elizabeth Wemmenhoven, Esther Meijer, Ilse Admiraal, Karlijn Pieterse, Kimberley van Vliet, Kimja Schouten, Linda Lorenz, Linette Thiessen, Marieke Graat, Nina Baker, Sanne Hofman, Sterre Rechtuijt, and Tjitske Dijkstra. We thank all individuals who agreed to post-mortem brain donation to the NBB, and their family members. We note that none of this work would be possible without the collaborative spirit and the exceptional dedication of the NBB staff, which explains that the NBB brain collection is of unprecedented high-quality.

## Inclusion & Ethics statement

All brain donors voluntarily agreed to postmortem brain donation and informed consent was obtained from all brain donors. The protocols of the 100-plus Study protocol and the Netherlands Brain Bank procedures were approved by the Medical Ethics Committee of the Amsterdam UMC. More information on ethics procedures can be found at https://www.brainbank.nl/about-us/ethics/.

## Author Contribution Statement

ABG, FK, KWL, AJMR and NBB collected and performed the proteomic measurements of the brain tissues donated to the 100-plus Study. MH, MZ, ABG, MJTR, FK and SSMM have performed the data analysis. MH, MZ, ABG, SSMM, JJMH, MJTR, ABS, and HH have written the manuscript. PS, JJMH, MJTR, ABS and HH supervised the research. HH designed the study. MH, MZ, ABG, FK, KWL, SSMM, ABS and HH have verified the underlying data. All authors read and approved the manuscript.

## Data Sharing Statement

The proteomics data is available in Table S2 and as an online resource at proteomics.holstegelab.eu.

## Material and Methods

### Cohorts

Tissues from Alzheimer’s disease (AD) patients and non-demented (ND) controls were selected from the brain cohort collected by the Netherlands Brain Bank (NBB, https://www.brainbank.nl/), and procedures are described in the Supplemental Data. In short, the NBB applied the Montine criteria^17^ or the neuropathologic assessment of AD patients: if the clinical report mentions a clinical diagnosis of dementia and we observe a high Braak stage for NFTs^74^ and high Thal stage for amyloid plaques^75^, the brain donor was considered an AD dementia case. In total, we included 61 ND controls spanning ages 50 to 96 and Braak stages 0 to III, and 91 AD patients with Braak stages from IV to VI, of which 48 AD patients had one copy of APOE-ɛ4 allele, 43 AD patients had no APOE-ɛ4 allele, spanning the ages 55 to 95 and 62 to 102, respectively. The average postmortem delay (PMD) ranges from 2.0 to 12.9 h (mean 5.7 h). Centenarian brains were donated to the 100-plus Study, a prospective cohort of centenarians in the Netherlands^8^. Inclusion criteria include self-reported and proxy-confirmed cognitive health and proof of age above 100 years. To objectively and thoroughly test cognitive health, all participants were visited yearly at their home and subjected to an extensive neuropsychological testing battery. Yearly visits continued until death or until participation was no longer possible. Around 30% of 100-plus Study participants agreed to post-mortem brain donation and tissue was collected in collaboration with the NBB (see Supplemental Data for procedures). At the time of tissue selection for the proteomics experiment, 58 centenarians aged 100 to 111 had come to autopsy and were included.

The average time between the last study visit and death is 9 months (±5 months) and postmortem delay ranges from 3.4 to 12.0 h (mean 6.5 h). Detailed information of all 210 brains analyzed in this study, their age, neuropathology stages and APOE genotype, the sex and PMD are listed in **Table S1**.

### Sample preparation

Fresh frozen tissue of the left middle temporal gyrus(MTG), anterior-posterior segment 2 of 5, was cut in 10 µm thick sections using a cryostat and mounted on polyethylene naphthalate-membrane slides (Leica, Herborn, DE). Sections were fixed in 100% ethanol for 1 minute and stained using 1% (wt/vol) toluidine blue in H2O (Fluka Analytical, Buchs, Switzerland) for 1 minute. Laser micro dissection (LMD) was performed using a Leica AS LMD system (Leica, Wetzlar, Germany) to isolate 0.5 mm^3^ of grey matter tissue and collected in 30 µl M-PER lysis buffer (Thermo Scientific, Rockford, IL, USA) in 0.5 ml Eppendorf PCR tubes and stored at -80 °C until further use.

Samples were heated to 95 °C for 5 minutes and incubated in the dark with 50 mM Iodoacetamide for 30 min at room temperature. Samples were loaded on 10% Bis/Tris polyacrylamide gels and run into the gel for 15 min at 80 V using 1.5 M Tris/Glycine SDS running buffer pH 8.3. Gels were fixed overnight and stained with colloidal Coomassie Blue G-250, before samples were cut out and small gel pieces of about 1 mm3 were placed in 96-well Nunc filter plates (Thermo Scientific, Rockford, IL, USA). Destaining, trypsin digestion, and peptide extraction were performed as described previously ^76^.

Collected samples were dissolved in 100 µl Mobile phase A (2% acetonitrile/0.1% formic acid) and cleaned using the OASIS filter plate (Waters Chromatography Europe BV, Etten-Leur, The Netherlands) according to the manufacturer’s instruction. We used a subset of all samples to generate a peptide library comprising 5 groups: (1) a pool of 4 young AD patients, (2) a pool of 4 young ND individuals, (3) a pool of 4 old AD patients, (4) a pool 4 old ND individuals, (5) a pool of 8 centenarians. We further fractionated the sample pools using the Pierce high-pH reversed phase fractionation spin columns (Thermo Scientific) according to manufacturer’s instruction but using 0.1% acetic acid instead of 0.1% trifluoroacetic acid. The collected peptides were dried and stored at -20 °C until mass spectrometry analysis.

## Mass Spectrometry

### Library generation

For the spectral library generation, we performed a data dependent acquisition (DDA) experiment using the five pooled samples (Sample preparation). Peptides were analyzed by micro LC MS/MS using an Ultimate 3000 LC system (Dionex, Thermo Scientific) coupled to the TripleTOF 5600 mass spectrometer (Sciex). Peptides were trapped on a 5 mm Pepmap 100 C18 column (300 μm i.d., 5 μm particle size, Dionex), and fractionated on a 200 mm Alltima C18 column (100 μm i.d., 3 μm particle size). The acetonitrile concentration in the mobile phase was increased from 5 to 18% in 88 min, to 25% at 98 min, 40% at 108 min and to 90% in 2 min, at a flow rate of 5 μl/min. The eluted peptides were electro-sprayed into the TripleTOF MS, with a micro-spray needle voltage of 5500 V. The mass spectrometer was operated in a data-dependent mode with a single MS full scan (350-1250 m/z, 150 msec) followed by a top 25 MS/MS (200–1800 m/z, 150 msec) at high sensitivity mode in UNIT resolution, precursor ion >150 counts/s, charge state from +2 to +5, with an exclusion time of 16 sec once the peptide was fragmented. Ions were fragmented in the collision cell using rolling collision energy, and a spread energy of 5 eV. The mass spectra were searched against the uniprot human fasta database (full proteome, release 2018-04) and iRT standard peptides using MaxQuant software (version 1.6.3.4) with default settings. The MaxQuant results were imported into Spectronaut (Biognosys) and converted into a spectral library.

### Sample analysis

Next, we measured the proteome of all 210 individuals, using data independent acquisition (DIA). The same LC gradient used by DDA was employed for DIA. The DIA protocol consisted of a parent ion scan of 150 ms followed by a selection window of 8 m/z with scan time of 80 ms, and stepped through the mass range between 450 and 770 m/z. The collision energy for each window was determined based on the appropriate collision energy for a 2+ ion, centered upon the window with a spread of 15 eV. The data were analyzed using Spectronaut with the default settings. Each group of eluting peptide fragments in the raw data was matched to the spectral library by Spectronaut and yielded a compound identification score for the assigned peptide. The false discovery rate (FDR) of this quality metric was provided in Spectronaut output as q-value. In total, 28,191 peptides from 4,829 unique proteins were measured in 210 proteomic profiles.

### Quality control

#### Sample filtering

The goal of sample filtering was to exclude low-quality proteomic profiles. High-quality samples were retained by removing those in which the proportion of low-quality peptides (q-value ≥ 0.01) exceeded 34%, corresponding to the point where the fraction of low-quality peptides increased sharply (n = 19; **Figure S1A,B**). In addition, samples with peptide abundance distributions deviating from the overall distribution (Kolmogorov–Smirnov distance > 0.04) were removed (n = 1; **Figure S1C,D**). After filtering, 190 proteome profiles remained for analysis: 88 AD patients, 53 controls, and 49 centenarians (Figure 1).

#### Peptide filtering

To ensure reliable protein abundance estimates, we filtered peptides based on quality metrics. For each protein, if ≥90% of measured samples contained at least one high-quality peptide (q-value < 0.01), the protein measurement was considered reliable (n = 3,448; **Table S2**); otherwise, it was excluded (n = 1,381). Protein abundances were log2-transformed prior to analysis.

Variance explanation analysis: To quantify sources of variation in protein expression, we applied a mixed-effect linear model using the variancePartition R package. The model estimated the proportion of variance explained by age, sex, Braak stage, NIA-Amyloid score, postmortem delay (PMD), APOE genotype, brain weight, and data acquisition batch. PMD, sex, and APOE genotype accounted for only minor proportions of variance. In contrast, batch effects explained a substantial share, alongside Braak stage and age, the main variables of interest (**Figure S2A**). Batch effects were subsequently corrected using the ComBat R package, after which reassessment confirmed that batch-related variance was effectively minimized (**Figure S2B**).

To further control for residual effects of PMD and sex, we constructed a regression model including sex, PMD, age, and group (AD, control, or centenarian) as predictors. Group and age were included due to their correlations with sex and PMD, respectively. Finally, the estimated contributions of sex and PMD were subtracted from the protein expression data.

### Neuropathological assessment

Neuropathological assessments of the NBB cohort and the 100-plus Study cohort were performed by the NBB, and thoroughly described the Supplemental Data. The centenarian brains and majority of the brains in the age-continuum were evaluated by a single neuropathologist, such that interrater variability was kept to a minimum.

### Quantitative immunohistochemistry (qIHC)

#### Phosphorylated Tau

To assess the load of phosphorylated tau, quantitative immunohistochemistry (qIHC) analysis with antibody AT8 was performed on the same MTG tissues on which proteomics was performed for a subset of postmortem brains: 75 AD cases, 35 ND controls and 20 centenarians (**Table S11**). The details of IHC staining and region of interest (ROI) identification are described in the Supplementary Material. Immunoreactive area (percentage) was determined using ImageJ software^77^.

For centenarians, Aβ load was determined in formalin-fixed paraffin-embedded sections of the right temporal pole (Brodmann area 38) as described previously^11^. In addition to general Aβ load (clone 6F/3D), we also quantified Aβ_40_ (clone G2-10) and Aβ_42_ (clone G2-11) specifically.

### Proteins associations

Associations were assessed using robust linear regression as implemented in the MASS package (v7.3-64). Statistical significance was evaluated using robust F-tests via the robftest function from the sfsmisc package (v1.1-20). P-values were adjusted for multiple testing using the False Discovery Rate (Benjamini–Hochberg procedure) with a significance threshold of 0.05.

Associations with AD case/control status, Braak stage, and NIA-Amyloid scores were analyzed within the combined AD and control groups, including age as an additional covariate. Models evaluating both amyloid and tau pathology were applied across all participant groups (AD, control, and centenarians) and included age, infarcts, TDP-43 amygdala, hippocampal sclerosis, and atherosclerosis as predictors. Atrophy and CERAD scores were excluded due to their strong correlation with AD pathology, and Braak Lewy Body stage was omitted owing to its low prevalence. Cerebral amyloid angiopathy (CAA) was included only in tau-specific analyses because of its correlation with amyloid pathology.

Models adjusted for cell-type composition incorporated four principal components derived from PCA summarizing variation in estimated cell fractions (see Cell type enrichment analysis).

Missing data were present for several pathology variables (TDP-43 amygdala: 30%, atrophy: 16%, atherosclerosis: 13%, CAA: 5%; lower for others; see **Table S1**). Missingness was addressed using multiple imputation (mice package, v3.16), including all pathology variables and age as predictors, with five imputations used to capture imputation uncertainty.

### Variance explained by pathology

To quantify the proportion of variance explained by different pathology measures, we constructed linear regression models using individual pathology scores as well as combinations thereof. For each model, residual variance after fitting was computed relative to the total variance in the dataset to assess explanatory power.

We next compared the performance of models based on NIA-Amyloid and Braak staging scores to those based on Aβ and tau peptide measurements for each protein. Standard linear regression models including age as a covariate were fitted for each comparison, and model fit was evaluated using the Akaike Information Criterion (AIC). Proteins showing a substantial difference in AIC (≥10) between models were identified and counted as strongly favoring one explanatory variable over the other.

As a sensitivity analysis, we explored several alternative modeling approaches: (i) merging Braak stages ≤3 into a single category to emphasize the marked pathological differences observed from stage ≥4 within the studied brain region, (ii) including additional pathology variables (CERAD, CAA, infarcts, hippocampal sclerosis, TDP-43 (amygdala), atrophy, atherosclerosis, and Braak (Lewy Body)) as supplementary predictors in both primary models; and (iii) applying polynomial regression models incorporating second- and third-degree terms of the assessed variables to evaluate potential non-linear effects.

### Pathway enrichment analysis

Pathway enrichment analysis was performed using Gene Set Enrichment Analysis (GSEA) implemented in the *WebGestaltR* package (v0.4.6)^78^. Multiple annotation resources were utilized, including Gene Ontology (GO) datasets (Biological Process, Cellular Component, and Molecular Function), pathway databases (KEGG, Panther, Reactome, and WikiPathways), the CORUM protein complex database, kinase target sets, transcription factor and miRNA target datasets, disease-related databases (GLAD4U, OMIM), drug-related databases (DrugBank, GLAD4U), and the Hallmark50 gene set collection.

Protein-level scores for GSEA were calculated as *–log₁₀(p-value) × sign(β)*. For peptides mapping to multiple proteins, only the first-listed protein was retained. Statistical significance was assessed via 10,000 permutations within *WebGestaltR*. Annotation terms were restricted to sets containing between 4 and 150 proteins.

To facilitate interpretation, overlapping annotation terms were grouped into clusters. Pairwise distances among annotation terms were computed using cosine distance, assigning a protein score of 0 to proteins absent from a given term and their corresponding score when present. Hierarchical clustering was performed using average linkage (implemented in *scipy* v1.15.2), and clusters were defined by cutting the resulting dendrogram at a cosine distance threshold of 0.4.

### Cell composition correction

Cell-type marker selection was performed as previously described^79^. In brief, since protein measurements were obtained from middle temporal lobe tissue, we integrated two single-cell datasets derived from the temporal cortex: (1) single-cell RNA-seq data from 466 cells across eight adult control donors^80^, and (2) single-nucleus RNA-seq data from 15,928 nuclei obtained from eight adult control donors^81^. These datasets were used to generate normalized gene expression profiles and corresponding cell-type label matrices. Expression-weighted cell type enrichment (EWCE) analysis was then conducted using the EWCE R package (version 1.2.0)^82^. The complete set of 3,448 proteins served as the background, from which 20,000 random lists were generated to bootstrap the probability distribution of cell-type expression. Proteins with cell specificity values ≥ 0.5 for a given cell type were classified as marker for that cell type.

Next, to estimate relative cell-type abundance, we first removed outlier markers for each cell type. For each marker, we computed its Pearson correlation with all other markers of the same type and iteratively excluded markers with median correlations below 0.2, starting from the lowest, until all remaining markers had median correlations greater than 0.2.

Using the remaining markers, we then derived a unified abundance profile per cell type through principal component analysis (PCA) on centered data. The first principal component captured the main pattern of coordinated abundance variation across markers and was interpreted as the primary measure of relative cell-type abundance. To maintain the original data scale when estimating cell-type fraction changes, we calculated a weighted average using the non-negative PCA loadings.

Because estimated abundance changes were strongly correlated among cell types, we performed an additional PCA across all estimated abundance components. For downstream association analyses that required correction for cell-type composition, we included the first four PCA components as covariates, which together accounted for 97.3% of the total variance in cell-type abundance.

### Synapse and mitochondrial marker proteins

To define protein marker sets for synaptic and mitchondrial structures (**Table S17, S18**), we used a two-stage procedure. First, candidate lists were generated with assistance from large language models (LLMs) to perform literature review. An initial list was generated using LLMs through an iterative querying process. Marker proteins were then organized according to their respective subcompartments, pathways, and complexes. To ensure accuracy and functional relevance, two additional LLMs were independently used to generate functional annotations for each proposed marker protein and to identify potential inconsistencies or errors.

Next, we validated all selected synaptic markers with SynGo^83^ and mitochondrial markers with MitoCarta 3.0^84^ annotations, by comparing these with LLM-based classifications. Based on this information, entries were manually validated through literature review to confirm biological plausibility and correctness.

### Protein abundance correlations

Correlations between data of the same modality (protein abundances or (sub) cell type abundances) were calculated using the Pearson correlation coefficient. Correlations between different modalities (e.g. protein abundances with neuropathological staging) were calculated using the Spearman rank correlation coefficient.

